# Cardiovascular complications are rare in severe COVID-19 presenting with myocardial injury

**DOI:** 10.64898/2026.01.04.26343417

**Authors:** Connor G O’Brien, Fatma Gunturkun, Wei Deng, Donald Chang, Sameer Khanijo, Christopher F Barnett, Shikha Kapil, Andrea Daly, Alberto Goffi, Dekel Stavi, François Martin Carrier, Yasmine Ennehas, Babar Fiza, Andrew Patterson, Thomas Daubert, Vanessa Moll, Clara Castellucci, Mitchell Zekhtser, Nibras Burgara, Michael Mayette, Karl Courchesne, Sean Thompson, Rushi Parikh, Juka S Kim, Melanie Ashland, Paul Mohabir

## Abstract

**Background:** SARS-COV2 infection has been linked to cardiovascular complications. Previously published data suggested that cardiac injury may be a common disease feature and contribute to morbidity and mortality in severe COVID-19. Since the early days of the pandemic, little data has emerged describing what cardiovascular complications arise from myocardial injury in severe COVID-19 and whether these complications contribute to mortality.

**Objectives:** Describe the incidence and nature of cardiovascular complications in patients with severe COVID-19 infection and biochemical or imaging evidence of myocardial injury.

**Methods:** We analyzed consecutive patients admitted for severe COVID-19 and presenting with biochemical or echocardiographic evidence of myocardial injury across 16 centers within the CARDIO-COVID investigator-initiated consortium. A light-GBM machine learning model was used to identify variables associated with mortality.

**Results:** Our study included 1,328 patients. In hospital mortality was 29.4%. Factors most associated with mortality were age, SOFA Cardiovascular score, elevated troponin, intubation, high vasoactive-inotropic score (VIS), high PEEP, and high FiO2. Cardiovascular complications were rare and did not present significant association with mortality. Patients with high VIS and SOFA Cardiovascular scores had a significantly higher incidence of secondary bacterial pneumonias suggesting septic shock played a role in their physiology. Investigator-adjudicated cause of death data identified respiratory failure and septic shock as primary causes of mortality.

**Conclusions:** Cardiovascular complications of severe COVID-19 are uncommon and do not seem to contribute significantly to mortality. Biochemical markers of myocardial injury or strain still carry significant prognostic value, but the value of these markers may indicate presence of high-risk ARDS phenotypes as opposed to significant myocardial injury.

## Introduction

The specter of impending cardiac failure has loomed over patients admitted with severe Coronavirus Disease 2019 (COVID-19) since the beginning of the pandemic [1]. Severe acute respiratory syndrome-related coronavirus 2 (SARS-CoV-2) is known to enter host cells via the angiotensin converting enzyme-2 (ACE2) receptor, which is heavily expressed in human myocardium[2]. Early reports of elevated troponin and NT-proBNP being predictive of severe COVID-19 raised concerns that myocardial injury played a significant role in severe disease [3–5]. Studies describing high rates of left ventricular dysfunction and various arrhythmias furthered this hypothesis, and contributed to the supposition of SARS-CoV-2 myocarditis as a common feature of COVID-19 [6–8].

Conflicting studies have emerged. Large cardiovascular databases suggest that cardiogenic shock and cardiovascular events are rare complications of COVID-19, yet the narrative supporting increased rates of myocarditis and death at the population level suggest that COVID-19 continues to have a large impact on cardiovascular health [9, 10]. Small cohort studies phenotyped patients suffering from acute cardiac failure due to severe COVID-19 showed multisystem inflammatory syndrome (MIS-A) as a cause of de novo cardiac failure [11], but patients suspected to have myocarditis without fitting this clinical syndrome rarely demonstrate histologic evidence to support the diagnosis [12]. Imaging studies have shown that ventricular dysfunction may contribute some predictive value for death but likely presents more as a sequelae of hypoxemia and elevated pulmonary vascular resistance than primary myocardial injury [13, 14].

The objective of this study was to investigate the frequency of cardiovascular complications in patients with severe COVID-19 and develop a predictive model describing how cardiovascular complications contribute to mortality. Contrary to earlier concerns regarding myocardial complications, we hypothesized that mortality among ICU patients with severe COVID-19 and evidence of myocardial involvement is primarily driven by respiratory complications, rather than cardiovascular complications.

## Methods

### Study Design and Patient Cohort

This is an ambidirectional multicenter cohort study conducted at 16 hospitals through a multi-national, investigator-initiated research network (CARDIO-COVID) of critical care physicians from the United States, Canada, Switzerland, and France. We included patients admitted to the ICU with COVID-19 and evidence of myocardial injury between April 2020 and April 2022. The data instrument was designed to define the cardiovascular effects of COVID-19 in critically ill patients with evidence of myocardial injury and predict how cardiovascular effects contribute to mortality. Myocardial injury was defined as a troponin above the upper limit of normal, BNP or NT-proBNP >10% the upper limit of normal, newly identified ejection fraction <50%, or a new drop in ejection fraction by >10% from baseline. The primary outcome was death during hospital admission. Data were collected both retrospectively and prospectively through comprehensive chart review and entered into a REDCap Database (Appendix 1). The CARDIO-COVID protocol and waiver of consent were approved by the Institutional Review Board at Stanford University and each of the participating centers. Stanford University School of Medicine served as the coordinating center. TRIPOD Statement principles were adhered to and checklist completed.

### Outcomes

Our primary outcome of interest was hospital mortality (or survival). Secondary outcomes were intubation, cardiac arrest, myocardial infarction, myocarditis, new onset heart failure, and stroke.

### Data collection

The data instrument collected patient demographics, comorbidities, cardiovascular complications (stroke, myocardial infarction, new onset heart failure, arrhythmia, cardiac arrest, myocarditis, and pulmonary embolism), infectious complications, and utilization data for advance ICU therapies including dialysis, intubation, and mechanical circulatory support. Serial data were collected for the first 15 days of ICU admission including: vital signs, labs, supplemental oxygen therapy, non-invasive positive pressure therapies, ventilator settings, and vasoactive medications and doses.

### Statistical Analysis

We summarized vital signs, labs and ICU resource utilization according to inpatient mortality. Due to a high degree of missingness within the cohort, a “missing data” category was established for all demographic and outcome variables. We employed descriptive statistics and absolute standardized differences (ASDs) to compare patients who died with those who did not. Categorical variables were presented as counts and percentages, while continuous variables were summarized as mean values along with their standard deviation (std). ASD measures of 0.2, 0.5, 0.8, and 1.3 corresponded to differences between the groups characterized as very small, small, medium, large, and very large, respectively.

Given broad dynamic ranges and low event rates at individual date points, continuous variables were dichotomized and high-risk categories defined based on the value that maximized the Youden’s J Statistic; peak FiO2 >60% for intubated, FiO2 >50% for non-intubated patients, troponin I >0.04ng/mL, troponin T >0.01ng/mL, and PEEP >10 cm H2O.

FiO2, troponin and PEEP had a high rate of missing data (Table 1). Instead of excluding these variables that could potentially influence mortality, we treated the missing data as a separate category and incorporated these variables into our prediction model. Given the high proportion of intubated patients, we differentiated higher risk lung injuries within that cohort. We determined threshold values for each variable by receiver operator curves (ROC) (Supplementary Figure 1).

**Table 1:** Depicts cause risk of death associated with demographic and clinical variables.

|  | Overall<br>(N=1328) | Died<br>(N=391) | Didn't Die<br>(N=937) | ASD |
| --- | --- | --- | --- | --- |
| **Age (years)** |  |  |  |  |
| Mean (SD) | 65 (15.6) | 69.6 (13.6) | 62.4 (16.1) | 0.478 |
| ≤ 65 | 459 (34.6%) | 116 (29.7%) | 343 (36.6%) | 0.448 |
| > 65 | 538 (40.5%) | 244 (62.4%) | 294 (31.4%) | 0.448 |
| Missing | 331 (24.9%) | 31 (7.9%) | 300 (32%) | 0.632 |
| **Sex** |  |  |  |  |
| Female | 473 (35.6%) | 154 (39.4%) | 319 (34%) | 0.144 |
| Male | 594 (44.7%) | 233 (59.6%) | 361 (38.5%) | 0.144 |
| **Race/Ethnicity** |  |  |  |  |
| Hispanic | 103 (7.8%) | 32 (8.2%) | 71 (7.6%) | 0.023 |
| Non-Hispanic American Indian/Alaskan Native | 2 (0.2%) | 0 (0%) | 2 (0.2%) | 0.065 |
| Non-Hispanic Asian | 51 (3.8%) | 31 (7.9%) | 20 (2.1%) | 0.267 |
| Non-Hispanic Native Hawaiian or Pacific Islander | 3 (0.2%) | 0 (0%) | 3 (0.3%) | 0.080 |
| Non-Hispanic Black or African American | 454 (34.2%) | 151 (38.6%) | 303 (32.3%) | 0.132 |
| Non-Hispanic White | 339 (25.5%) | 119 (30.4%) | 220 (23.5%) | 0.157 |
| Unknown / Not Reported | 376 (28.3%) | 58 (14.8%) | 318 (33.9%) | 0.456 |
| **Medical History** |  |  |  |  |
| Hypertension | 767 (57.8%) | 279 (71.4%) | 488 (52.1%) | 0.405 |
| Diabetes | 517 (38.9%) | 204 (52.2%) | 313 (33.4%) | 0.386 |
| Asthma | 60 (4.5%) | 25 (6.4%) | 35 (3.7%) | 0.121 |
| Chronic obstructive pulmonary disease | 119 (9%) | 54 (13.8%) | 65 (6.9%) | 0.227 |
| Chronic lung disease | 43 (3.2%) | 22 (5.6%) | 21 (2.2%) | 0.175 |
| Chronic kidney disease | 269 (20.3%) | 96 (24.6%) | 173 (18.5%) | 0.149 |
| Chronic dialysis | 59 (4.4%) | 23 (5.9%) | 36 (3.8%) | 0.095 |
| Coronary artery disease | 183 (13.8%) | 82 (21%) | 101 (10.8%) | 0.282 |
| Solid organ transplant | 28 (2.1%) | 13 (3.3%) | 15 (1.6%) | 0.111 |
| Heart failure with preserved ejection fraction | 97 (7.3%) | 39 (10%) | 58 (6.2%) | 0.139 |
| Heart failure with reduced ejection fraction | 64 (4.8%) | 31 (7.9%) | 33 (3.5%) | 0.191 |
| Bone marrow transplant | 2 (0.2%) | 2 (0.5%) | 0 (0%) | 0.101 |
| LVAD implantation | 3 (0.2%) | 1 (0.3%) | 2 (0.2%) | 0.009 |
| Atrial fibrillation | 175 (13.2%) | 68 (17.4%) | 107 (11.4%) | 0.171 |
| Severe cardiac valvular disease | 31 (2.3%) | 12 (3.1%) | 19 (2%) | 0.066 |
| Pulmonary hypertension diagnosis | 29 (2.2%) | 10 (2.6%) | 19 (2%) | 0.035 |
| Malignancy | 83 (6.2%) | 43 (11%) | 40 (4.3%) | 0.255 |
| None of the above | 108 (8.1%) | 30 (7.7%) | 78 (8.3%) | 0.024 |
| **Outcomes** |  |  |  |  |
| Myocarditis | 45 (3.4%) | 27 (6.9%) | 18 (1.9%) | 0.244 |
| Stroke | 44 (3.3%) | 19 (4.9%) | 25 (2.7%) | 0.115 |
| Intubation | 497 (37.4%) | 290 (74.2%) | 207 (22.1%) | 1.221 |
| STEMI | 27 (2%) | 17 (4.3%) | 10 (1.1%) | 0.203 |
| New Heart Failure | 55 (4.1%) | 29 (7.4%) | 26 (2.8%) | 0.212 |
| Cardiac Arrest | 54 (4.1%) | 51 (13%) | 3 (0.3%) | 0.527 |
| Chest Compressions | 47 (3.5%) | 39 (10%) | 8 (0.9%) | 0.394 |
| Pneumonia | 182 (13.7%) | 106 (27.1%) | 76 (8.1%) | 0.515 |
| Torsades de Pointes / Sustained Ventricular Tachycardia | 44 (3.3%) | 27 (6.9%) | 17 (1.8%) | 0.251 |
| Atrial Fibrillation / Atrial Flutter / Sustained Supraventricular Tachycardia | 211 (15.9%) | 117 (29.9%) | 94 (10%) | 0.514 |
| Bradycardia requiring therapy | 37 (2.8%) | 22 (5.6%) | 15 (1.6%) | 0.217 |
| **VIS Max** |  |  |  |  |
| VIS > 0: Mean (SD) | 193.1 (218.5) | 248.7 (225.4) | 110.6 (178.8) | 0.681 |
| VIS = 0 | 573 (43.1%) | 104 (26.6%) | 469 (50.1%) | 0.987 |
| **SOFA Cardiovascular** |  |  |  |  |
| 0 | 508 (38.3%) | 71 (18.2%) | 437 (46.6%) | 1.166 |
| 1 | 14 (1.1%) | 9 (2.3%) | 5 (0.5%) | 0.137 |
| 2 | 4 (0.3%) | 1 (0.3%) | 3 (0.3%) | 0.032 |
| 3 | 234 (17.6%) | 124 (31.7%) | 110 (11.7%) | 0.407 |
| 4 | 212 (16%) | 146 (37.3%) | 66 (7%) | 0.753 |
| Missing | 356 (26.8%) | 40 (10.2%) | 316 (33.7%) | 0.592 |
| **Troponin Max** |  |  |  |  |
| Normal | 74 (5.6%) | 23 (5.9%) | 51 (5.4%) | 0.320 |
| High Risk | 416 (31.3%) | 221 (56.5%) | 195 (20.8%) | 0.320 |
| Missing | 838 (63.1%) | 147 (37.6%) | 691 (73.7%) | 0.781 |
| **FIO2 Max** |  |  |  |  |
| Normal | 96 (7.2%) | 22 (5.6%) | 74 (7.9%) | 0.599 |
| High Risk | 355 (26.7%) | 206 (52.7%) | 149 (15.9%) | 0.599 |
| Missing | 877 (66%) | 163 (41.7%) | 714 (76.2%) | 0.749 |
| **PEEP Max (in Intubated or BiPAP patients)** |  |  |  |  |
| > 10 | 174 (13.1%) | 109 (27.9%) | 65 (6.9%) | 0.016 |
| ≤ 10 | 93 (7%) | 59 (15.1%) | 34 (3.6%) | 0.016 |

To assess the predictive capacity of clinical characteristics for mortality, we employed 23 clinical features including age, Vasoactive-Inotropic Score (VIS), Sequential Organ Failure Assessment (SOFA) cardiovascular score, Troponin, Peep, and FiO2. A Light Gradient Boosting Machine (LightGBM) model was developed using these features. The dataset was partitioned into training (60%), validation (20%), and testing (20%) subsets. Hyperparameter tuning was performed on the validation dataset using Bayesian optimization. Model performance was subsequently evaluated on the test dataset, considering sensitivity, specificity, and the area under the curve (AUC) with two-sided 95% confidence intervals (CIs). Furthermore, we utilized the Shapley additive explanation (SHAP) method to assess the individual impact of each feature on the likelihood of mortality. A SHAP value closer to 0 means that no relationship can be established between a variable and outcome of interest while values closer 1 suggest a stronger relationship. Patients with new onset heart failure were analyzed separately to assess whether systemic complications of cardiac failure bore a greater relationship with mortality in this population. SHAP was further utilized to assess in detail the factors and their interactions that are most associated with mortality. All analyses were performed using Python and R programming languages.

## Results

A total of 1328 patients were entered into the CARDIO-COVID registry, among whom 391 (29.4%) experienced mortality. The mean (std) age was 65 (15.6) years, with 594 (44.7%) being male, and 103 (7.8%) identifying as Hispanic. The predominant comorbidities were hypertension (57.8%), diabetes (38.9%) and chronic kidney diseases (20.3%). (**Table 1**). Within the cohort 497 (37.4%) patients required intubation, 55 (4.1%) experienced new onset heart failure, 54 (4.1%) experienced cardiac arrest, 45 (3.4%) were thought to have myocarditis, 44 (3.3%) experienced a stroke, 27 (2.7%) experienced STEMI. Distribution of patients by center is shown in Supplementary Table 1.

Cause of death data further supported respiratory failure and hypoxemia as the leading cause of mortality (**Figure 1**). Cause of death in rank order were hypoxemic respiratory failure (N = 106, 65.4%), septic shock (N = 17, 10.5%), cardiac arrest due to Pulseless Electrical Activity (PEA) (N = 15, 9.3%), cardiac failure (N = 5, 3.1%), cerebrovascular accident (N = 3, 1.9%), and hemorrhagic shock (N = 2, 1.2%). Of patients who died of cardiovascular complications: 1 died of an NSTEMI, 1 died of bradycardia, 1 died of cardiogenic shock, and two experienced cardiogenic shock in the context of multiorgan failure.

**Figure 1:**
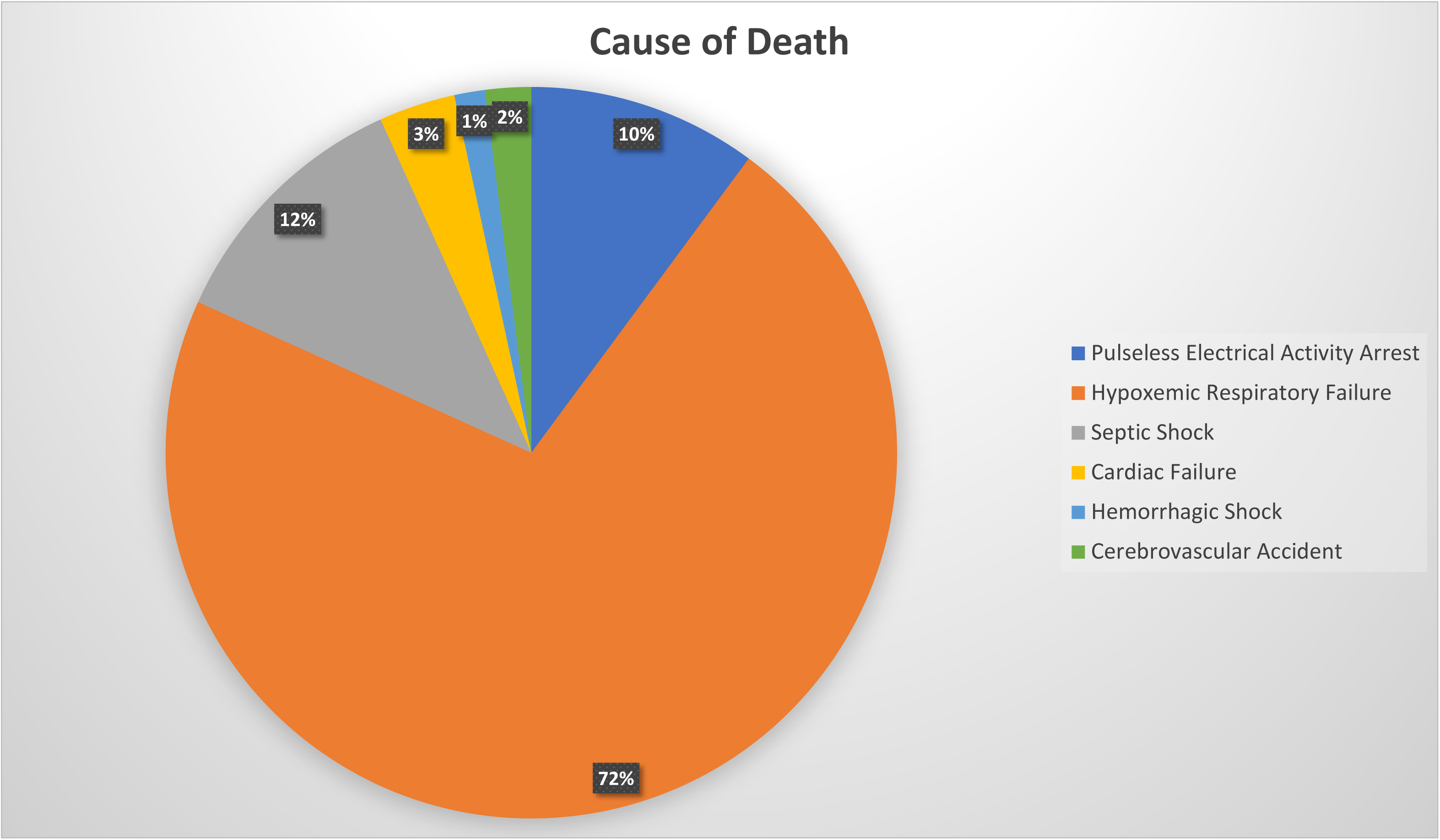
Depicts cause of death (N = 391) as adjudicated by site investigators.

Table 1 presents descriptive statistics of patients who experienced mortality, in comparison with those who survived. As anticipated, the proportion of older patients was higher in the mortality group compared to those who survived (62.4% vs. 31.4%, ASD=0.45). Consistent with previous findings [7, 15], the mortality group demonstrated an elevated prevalence of comorbid conditions such as hypertension (71.4% vs. 52.1%, ASD: O.41), diabetes (52.2% vs. 33.4%, ASD:0.39) and coronary artery disease (21% vs. 10.8%, ASD:0.28). On the contrary, differences in male patients (63.2%, vs. 53.1%, ASD:0.14) or those with atrial fibrillation (17.4% vs. 11.4%, ASD:0.17) and chronic kidney disease (24.6% vs. 18.5%, ASD:0.15) were small.

Of the outcomes, intubation (74.2% vs. 22.1%, ASD:1.22), concomitant bacterial pneumonia (27.1% vs. 8.1%, ASD:0.52), and new onset a-fib (29.9% vs. 10%, ASD:0.51) were both common in the mortality group. Stroke, myocardial infarction, ventricular dysrhythmias, pulmonary embolism, myocarditis and new onset heart failure were uncommon and presented similar incidences in both survivors and non-survivors.

Of the clinical variables, maximum VIS score, SOFA Cardiovascular Score, PEEP, FiO2, and serum troponin demonstrated medium to large differences between the mortality and surviving groups. PEEP >10 cm H2O showed the largest difference between the groups (27.9% vs 6.9%, ASD 0.86). Similarly, high risk category in FiO2 demonstrated a large difference (52.7% vs 15.9%, ASD 0.60), while combined troponin T and I above cutoff showed a medium difference between the groups (56.5% vs 20.8%, ASD 0.32).

Our LightGBM model was tested on 266 patients, with 78 (29.3%) patients experiencing mortality. The model achieved an accuracy of 80.1%, sensitivity of 78.2%, specificity of 80.9%, and an AUC of 0.89 (95% CI, 0.85-0.93) (**Figure 2**). The most important variable predicting mortality was age, followed by higher SOFA Cardiovascular, need for intubation, and elevated troponin (**Figure 3**). The presence of PEA had a strong relationship with mortality, but the absence was not protective, which is why its association with mortality was less than the aforementioned variables. The likelihood of mortality increased with higher values of the respective variables. Importantly, cardiovascular complications including new onset heart failure, ventricular tachycardia/fibrillation, and suspected myocarditis showed very little predictive value for mortality (**Figure 3**).

**Figure 2:**
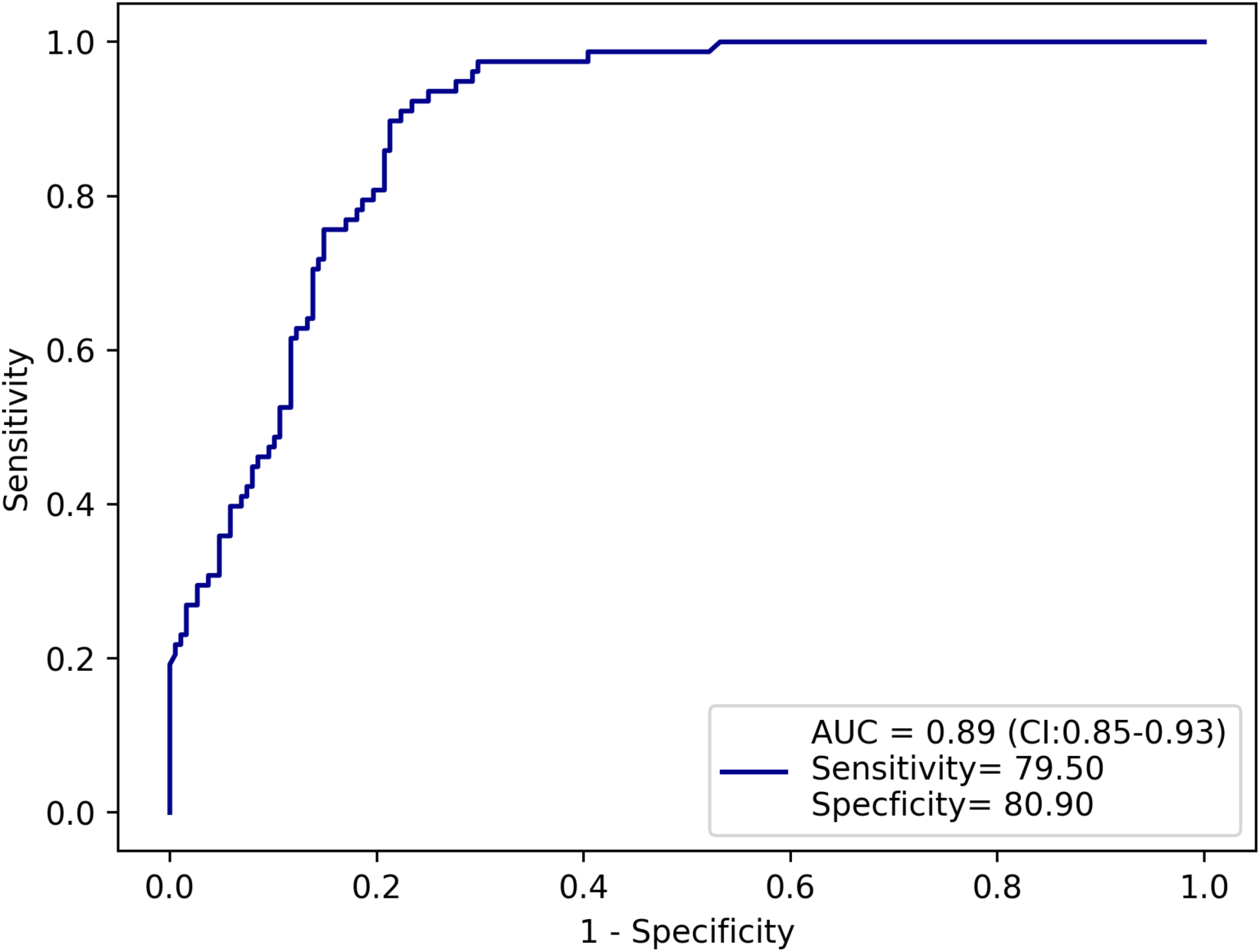
Depicts sensitivity and specificity of the Light GBM model. Area Under the Curve (AUC) displayed in the bottom right-hand corner.

**Figure 3:**
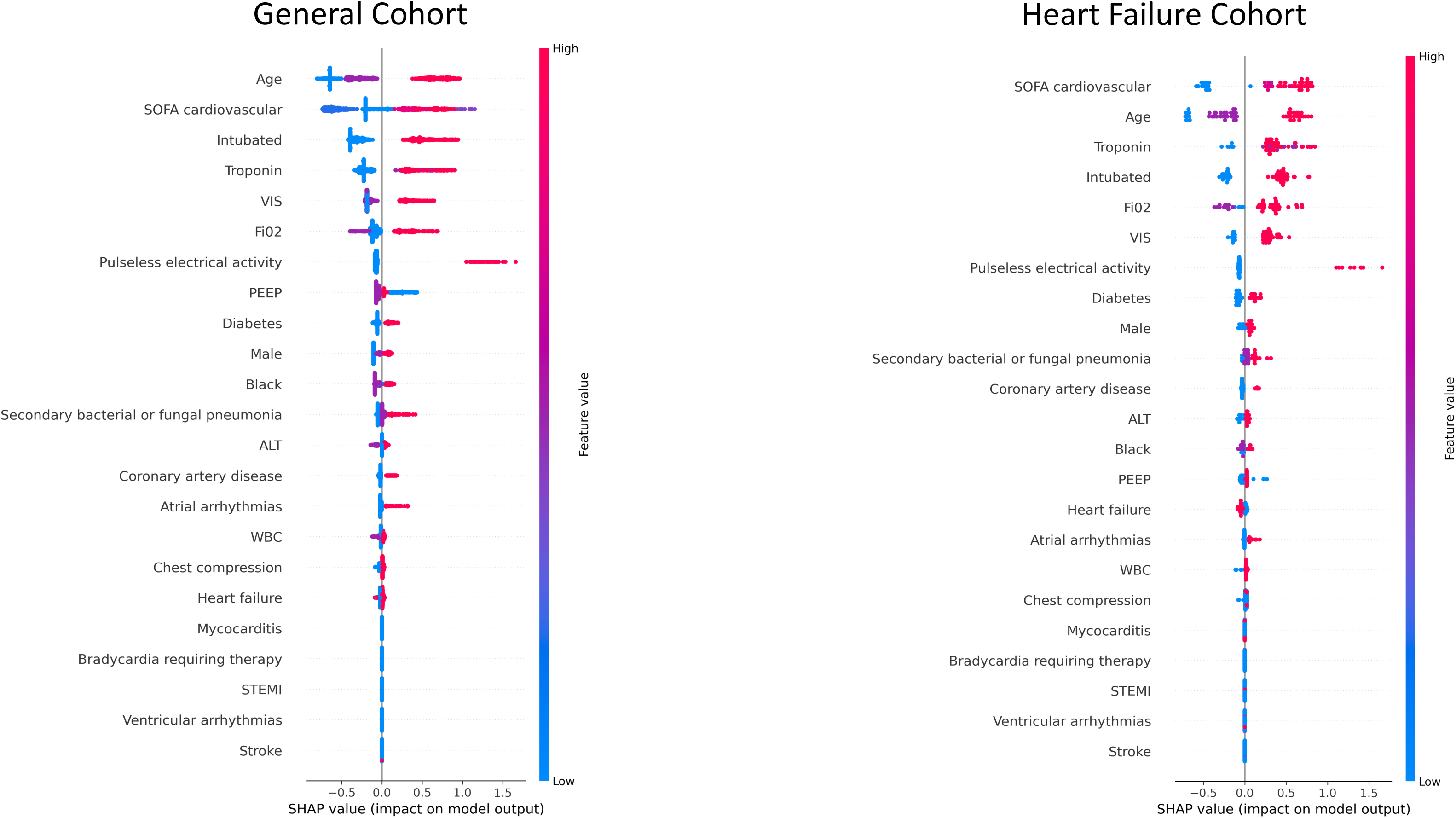
Graphical depiction of relative impact each clinical variable had on risk of death in the light GBM model. Data elements at the top carried the greatest association with mortality. The entire CARDIO COVID cohort shown on the left and heart failure cohort on the right. Each hash mark represents an individual patient.

To achieve better insights into how hemodynamic compromise and lung injury relate to one another with respect to risk of death we performed 3D SHAP plots plotting three variables simultaneously, which can be seen in **Figure 4**. When separated by age >65 or <65 (**Figure 4a**), older patients who were not intubated had a higher risk of death despite having lower SOFA Cardiovascular score. Within patients older than 65, non-intubated patients who died had a mean age of 79 compared to 76 for intubated patients who died (p = 0.003).

**Figure 4:**
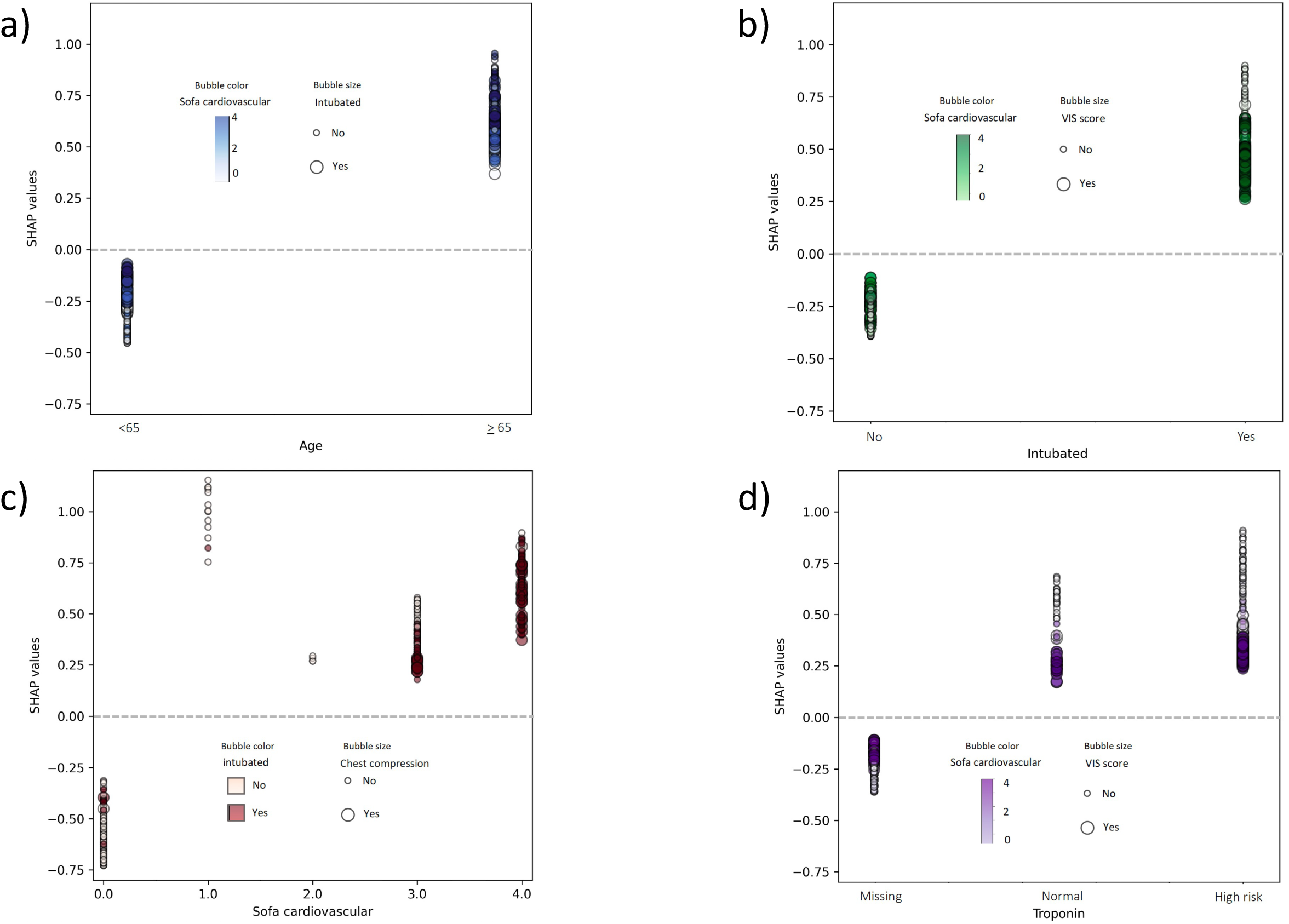
Depicts contribution of high-risk variables to individual patient SHAP value. Color and dot size reflect additional high-risk variables describing mechanism of injury associated with variable in question. Dots represent individual patients. a) When patients analyzed by age, even distribution of SOFA Cardiovascular scores and intubation status demonstrates that age carries the greatest risk of death. b) Analyzing by intubated status highlights the population with likely isolated viral pneumonia and low SOFA Cardiovascular and VIS scores. c) When analyzed by SOFA Cardiovascular score, increasing risk and complications of hemodynamic compromise likely driven by septic shock and chest compressions is demonstrated. d) Analysis by troponin level demonstrates increasing SHAP values but even distribution of indicators of hemodynamic compromise suggesting troponin does not implicate cardiac failure.

When patients were segregated by intubation status, SOFA Cardiovascular, and VIS (**Figure 4b**), a separation appears where the patients at greatest risk for mortality were intubated patients with low SOFA Cardiovascular and VIS scores. This may reflect the presence of a treatable process like secondary bacterial pneumonia and septic shock driving respiratory decompensation in patients with higher SOFA scores while intubated patients with low SOFA scores succumbed to irreversible viral lung injury without sepsis. In the intubated cohort bacterial pneumonia was reported more in patients with a SOFA Cardiovascular score >1 in 25.8% vs 5.3% in those with SOFA score <1 (ASD: 0.59).

When patients are segregated by SOFA Cardiovascular score and then analyzed by exposure to intubation and chest compressions we see further separation of populations. **Figure 4c** shows that patients with higher SOFA Cardiovascular scores, greater exposure to intubation, and exposure to chest compressions carried the highest risk for mortality. Patients with a SOFA score greater than 1 had a significantly higher risk of pneumonia suggesting septic shock contributed to higher scores (5.3% vs 24.9%, p = 0.0001, Supplementary Table 2). Again, the elderly, non-intubated cohort at high risk for mortality is shown as an outlier population in the SOFA Cardiovascular score of 1 category.

In **Figure 4d** patients are segregated by high-risk or low-risk troponin status; serum troponin above the upper limited of normal were associated with mortality. Within this cohort, the act of checking a troponin carried predictive value for death. The average troponin and dynamic range were narrow precluding clinically meaningful evaluation of troponin as a continuous variable.

Given low event rates, we were unable to analyze outcomes specifically within populations of patients who experienced cardiovascular complications except for patients with newly identified heart failure with reduced ejection fraction (HFrEF). Within this population the mortality rate was 54.3% (38/70). The higher mortality rate in this cohort was most attributable to age. The mean (std) age for HFrEF patients who died was older [67.5(12.9), ASD: 0.71], compared to patients who survived [55.2(17.2)]. Ejection fraction did not differ significantly between survivors and non-survivors (39.4 (10.2) vs 37.4 (12.7), p = 0.7).

Within the heart failure population predictors of death were similar to the overall cohort with the exception of SOFA Cardiovascular and VIS scores (Table 2). Absolute VIS scores were higher within the heart failure population compared to the general cohort. Light GBM analysis similarly demonstrates that elevated SOFA Cardiovascular and VIS scores carry greater predictive value for death in the heart failure cohort than the general cohort (Figure 3). Secondary bacterial or fungal pneumonia was more common in heart failure patients with SOFA Cardiovascular scores >1 suggesting septic shock may be contributing to increased SOFA Cardiovascular and VIS scores (Supplementary Table 3).

**Table 2:**
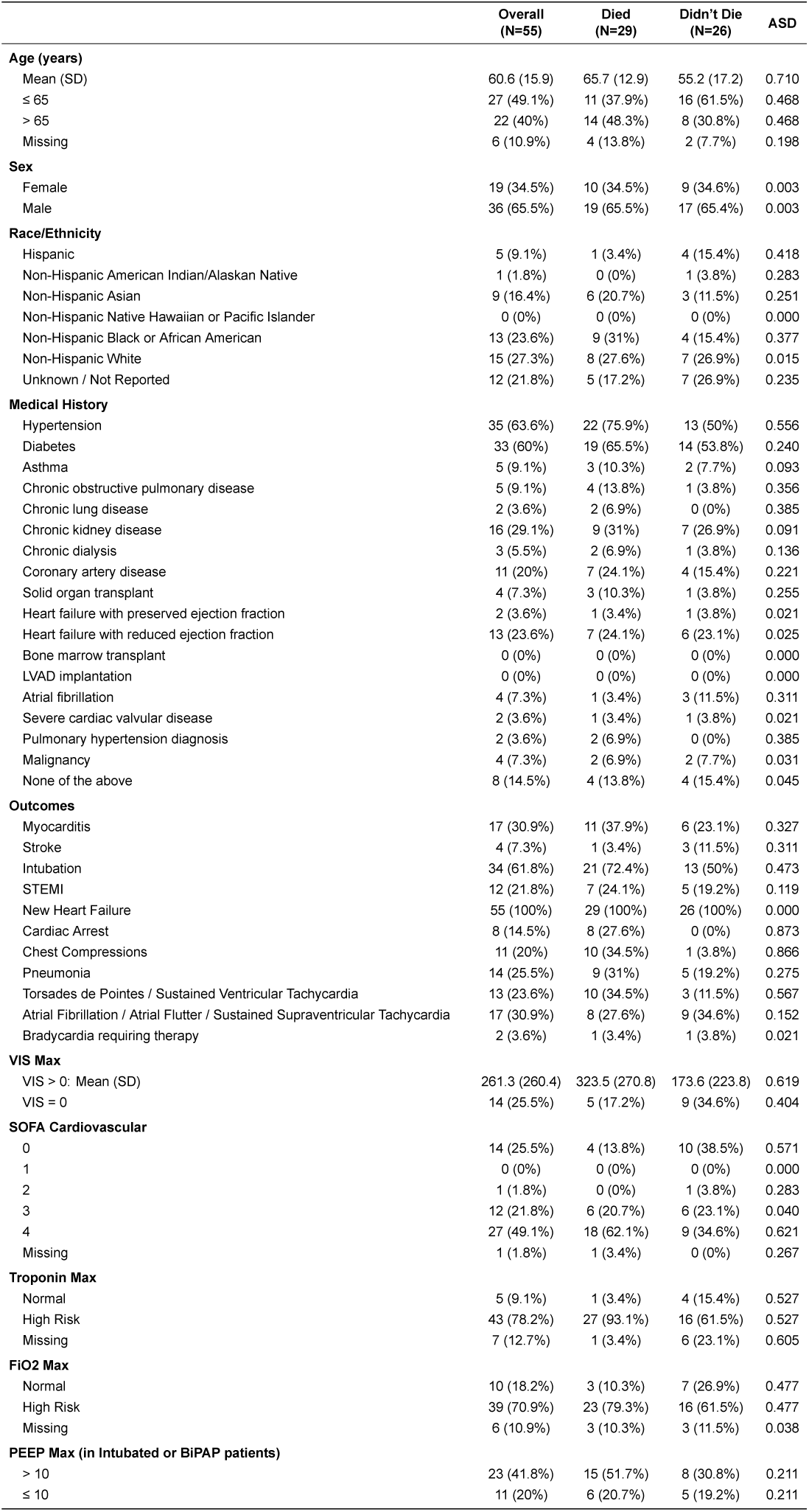
Depicts risk of death associated with demographic and clinical variables for patients who experienced new onset heart failure.

## Discussion

Our analysis of the CARDIO-COVID registry demonstrates that cardiovascular events are infrequent in severe COVID-19 disease and not significantly associated with mortality (Central Figure). By enriching our population for patients with lab evidence of myocardial injury and/or echocardiographic evidence of new myocardial dysfunction, we intended to select for patients most likely to be harboring a cardiovascular insult. Instead, the CARDIO-COVID cohort demonstrates that respiratory failure, cardiac arrest due to PEA, and septic shock are the primary causes of death.

Consistent with prior cohorts, our population shows that elderly patients and those with risk factors for cardiovascular disease are at the highest risk for death. Troponin continues to demonstrate an association with poor outcome but not cardiovascular events. Instead, elevated troponin is associated with increased risk of severe lung injury, and degree of lung injury does not correlate with degree of troponin elevation. Similarly, high SOFA Cardiovascular and VIS scores carry an increased risk of death in both the general and heart failure cohorts, but the majority of these patients died from septic shock or hypoxemic respiratory failure. Rates of secondary bacterial pneumonia were significantly higher in patients with SOFA Cardiovascular scores >1 in both the general and heart failure cohorts suggesting sepsis as the predominant cause for hemodynamic compromise.

Cardiovascular complications of COVID-19 garnered significant attention throughout the pandemic. SARS-CoV2 virus enters cells via the angiotensin receptor [2], and early reports described troponin positivity being associated with poor outcomes suggesting that myocardial injury may be a source of pathogenesis [1, 4]. These reports were followed by studies from selected populations reporting increased risk of cardiovascular complications including heart failure [16], arrhythmia [7, 17], and cardiac arrest [18] each being associated with increased risk of death. Studies from large, unselected populations failed to recapitulate the same risks of cardiovascular complications [5, 9] and, conversely, demonstrate a low incidence (<1%) of death attributable to cardiovascular causes [19]. Results from CARDIO-COVID, despite being a population of patients thought to be enriched for cardiovascular complications, reflect similarly low individual rates of cardiovascular complications, low incidence of cardiovascular death, high incidence of non-cardiogenic shock, and high incidence of all-cause mortality. Unique to this study, cause of death data was adjudicated by site investigators directly involved in patient care during the pandemic, and their observations corroborated our LightGBM model demonstrating that the overwhelming causes of death were respiratory failure and septic shock.

Given the low event rate, a LightGBM model is particularly well suited to analyze the impact of cardiovascular events [20]. LightGBM decision trees efficiently identify high magnitude but low frequency events, distinct high risk subgroups, and non-linear risk relationships [21]. We show that cardiovascular events are not only infrequent but do not carry outsized risk of death. Our model demonstrated high accuracy, sensitivity, and specificity when using age, intubation, SOFA Cardiovascular score, and troponin to predict ICU mortality. Our model identified PEA as a rare but high-risk event then appropriately weighted PEA lower given the absence of PEA was not protective for survival. We did not see a similar effect for any cardiovascular complications.

While our findings describe low risks of cardiovascular complications of COVID-19, rare cases of acute, fulminant heart failure due to myocarditis have been described [11]. Though myocarditis can be catastrophic, incidence of myocarditis in COVID-19 is <0.15% in hospitalized patients [22] and the majority of these patients do not experience hemodynamic insults. It is also important to note that degree of troponin and naturetic peptide elevation are not dissimilar between severe myocarditis associated with heart failure and patients with severe lung COVID-19 lung disease.

Cardiac biomarkers have been recognized as valuable indicators of critical illness. Severe systemic inflammation from sepsis and exacerbations of chronic lung disease have been shown to be associated with significant elevations in troponin and naturiuretic peptide when cardiac function and filling pressures are normal [23, 24]. Inflammatory cytokines and proteins have been used to phenotype acute respiratory distress syndrome (ARDS), both offering insights into disease mortality and novel treatment opportunities [25]. In severe COVID-19 troponin has more recently been recognized as an indicator of hyperinflammatory ARDS, a disease process that that responds more favorably to corticosteroids than hypoinflammatory ARDS phenotypes [26]. Recognizing elevated cardiac biomarkers may indicate hyperinflammatory lung injury could both expedite appropriate treatment of ARDS and avoid unnecessary cardiac diagnostics.

The primary limitations of the CARDIO-COVID registry are the retrospective nature of the data collection as well as the high degree of missingness. While retrospective data carries opportunities for unrecognized bias, the infrequent nature of cardiovascular events in COVID makes prospective data collection challenging. We selected for patients at greatest risk for cardiac complications to minimize the chance that cardiovascular complications are underrepresented.

The high degree of missingness is explained by the most affected variables coming from the serial vital signs sheets, which were not required entries. Furthermore, handling of patients with missing variables as an independent group did not affect relationships between demographics, comorbidities, or use of ICU therapies and mortality or mode of death.

Given the investigator-initiated nature of the study and stressed working conditions of the pandemic, missing data is a common challenge. By analyzing missingness as an independent variable we showed that within this dataset missingness does not appear to bias associations. Missingness was most common in the daily ICU flow sheet data form, which was an optional data field for investigators. Outcomes and cause of death data had a high rate of completeness rendering the low cardiovascular event rates and associations likely valid. These findings are similar to studies using from inpatient datasets not enriched for cardiovascular involvement[19].

The primary strengths of the registry are the multinational structure, enriched patient population, duration of the study, and investigator driven data entry. The multinational structure and duration of the study allow for the associations to describe outcomes for multiple SARS CoV-2 variants. Data entry was performed by investigators with direct knowledge of patient care and outcomes. This allowed for a high degree of data fidelity and collection of cause of death.

### Clinical Perspective

Cardiovascular events are less significant drivers of mortality in severe COVID-19 than previously described. In a population thought to be at high risk for cardiovascular death based on early data published in the COVID-19 Pandemic, the overwhelming causes of mortality were hypoxemic respiratory failure and septic shock[27, 28]. Biomarkers of cardiac injury may represent a hyperinflammatory ARDS phenotype rather than significant myocardial injury in the majority of cases[29].

**Central Figure:**
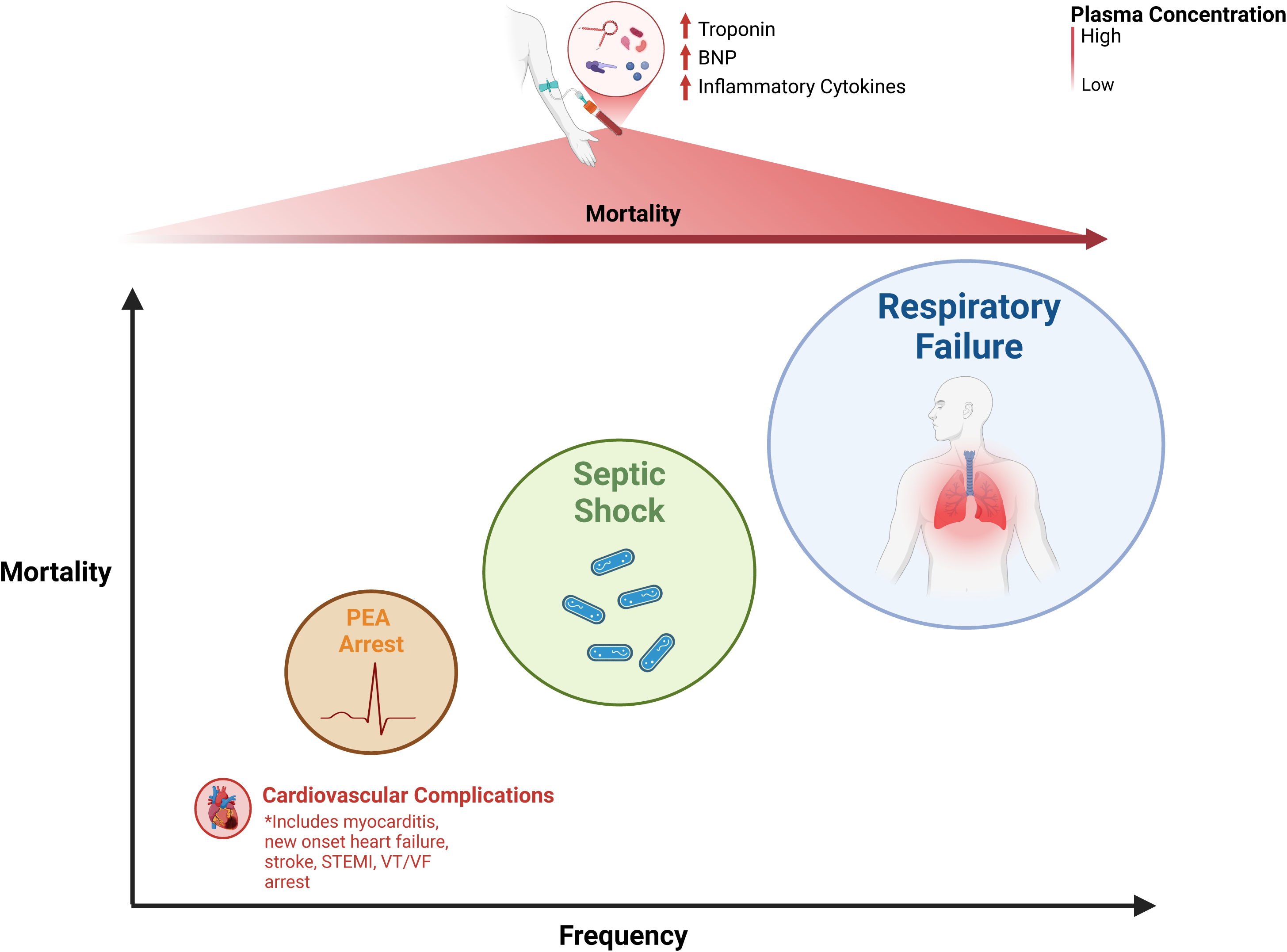
Patients admitted to ICUs with severe COVID-19 and evidence of myocardial injury (top) most commonly die from sequelae of acute respiratory distress syndrome, i.e. respiratory failure, septic shock, and cardiac arrest mediated by prolonged hypoxemia. Despite every patient demonstrating evidence of myocardial injury in the setting of severe COVID-19, cardiovascular complications occur in ∼15% of patients (mostly atrial fibrillation), and lead to patient death in 3% of cases (bottom left). Created with BioRender.com.

## Data Availability

All data produced in the present study are available upon reasonable request to the authors.

## Supplementary Tables and Figures

**Supplementary Figure 1:**
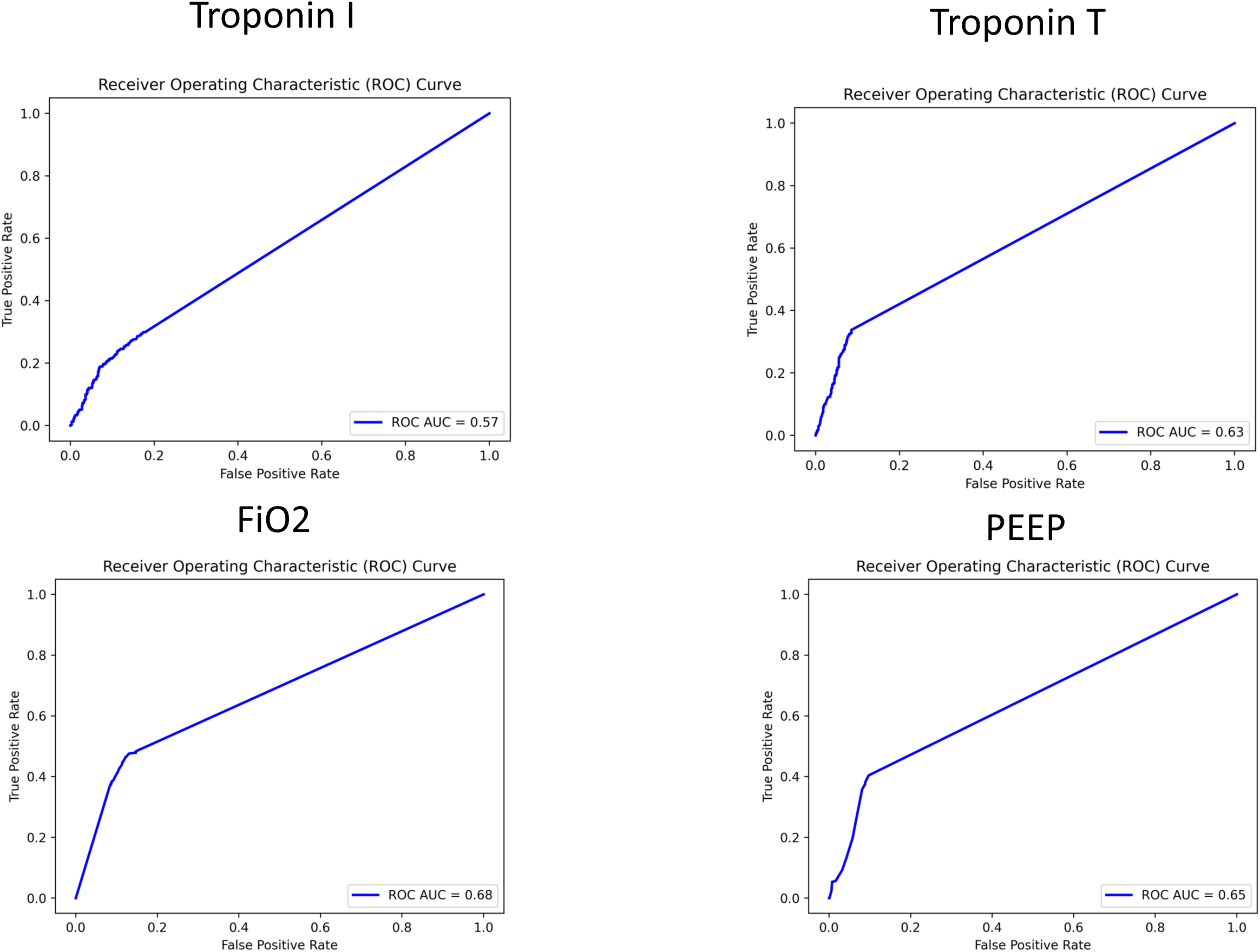
Receiver operator curves showing AUCs and thresholds associated with high-risk for mortality

**Supplementary Table 1:** Patients entered per center.

| Participating Center | Patients Entered |
| --- | --- |
| Ochsner Medical Center | 641 |
| Emory University | 128 |
| Medstar Medical Center | 99 |
| University of Toronto | 92 |
| Stanford University | 80 |
| Feinstein Institute of Medical Research | 79 |
| Centre Hospitalier de L'Université de Montréal | 51 |
| University of Pennsylvania | 35 |
| University of Zurich | 26 |
| Université de Sherbrooke | 23 |
| University of California Los Angeles | 21 |
| University of Albany | 14 |
| Univesity of Nebraska | 13 |
| Wake Forest Univeristy | 5 |
| Clinique Ambroise Paré Neuilly | 2 |
| Massachusetts General Hospital | 1 |

**Supplementary Table 2:**
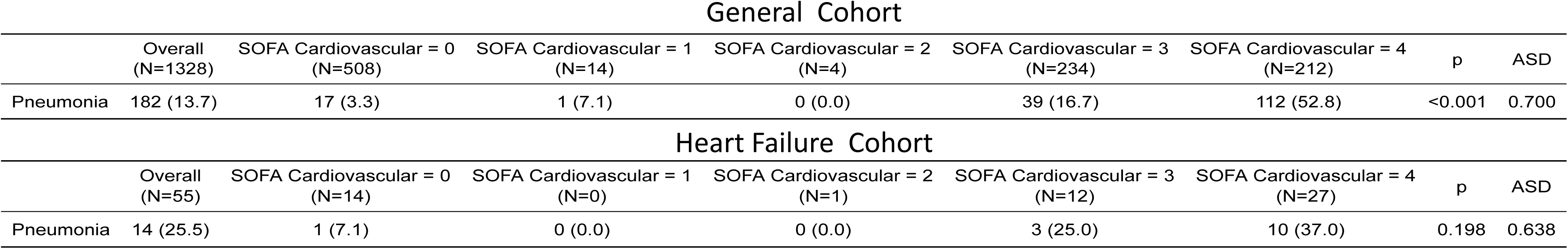
Patients diagnosed with secondary bacterial or fungal pneumonia analyzed by SOFA Cardiovascular score. Separation of patients demonstrates risk of greater hemodynamic compromise in patients diagnosed with pneumonia reflecting increased risk of septic shock.

General Cohort
|  | Overall<br>(N=1328) | SOFA Cardiovascular = 0<br>(N=508) | SOFA Cardiovascular = 1<br>(N=14) | SOFA Cardiovascular = 2<br>(N=4) | SOFA Cardiovascular = 3<br>(N=234) | SOFA Cardiovascular = 4<br>(N=212) | p | ASD |
| --- | --- | --- | --- | --- | --- | --- | --- | --- |
| Pneumonia | 182 (13.7) | 17 (3.3) | 1 (7.1) | 0 (0.0) | 39 (16.7) | 112 (52.8) | <0.001 | 0.700 |

|  | Overall<br>(N=55) | SOFA Cardiovascular = 0<br>(N=14) | SOFA Cardiovascular = 1<br>(N=0) | SOFA Cardiovascular = 2<br>(N=1) | SOFA Cardiovascular = 3<br>(N=12) | SOFA Cardiovascular = 4<br>(N=27) | p | ASD |
| --- | --- | --- | --- | --- | --- | --- | --- | --- |
| Pneumonia | 14 (25.5) | 1 (7.1) | 0 (0.0) | 0 (0.0) | 3 (25.0) | 10 (37.0) | 0.198 | 0.638 |

## Notes

### Competing Interest Statement

The authors have declared no competing interest.

### Funding Statement

This study did not receive any funding.

### Author Declarations

The IRB of Stanford University gave ethical approval for this work.

