## Supplementary Data for "Cardiovascular complications are rare in severe COVID-19 presenting with myocardial injury"

### Slide 1
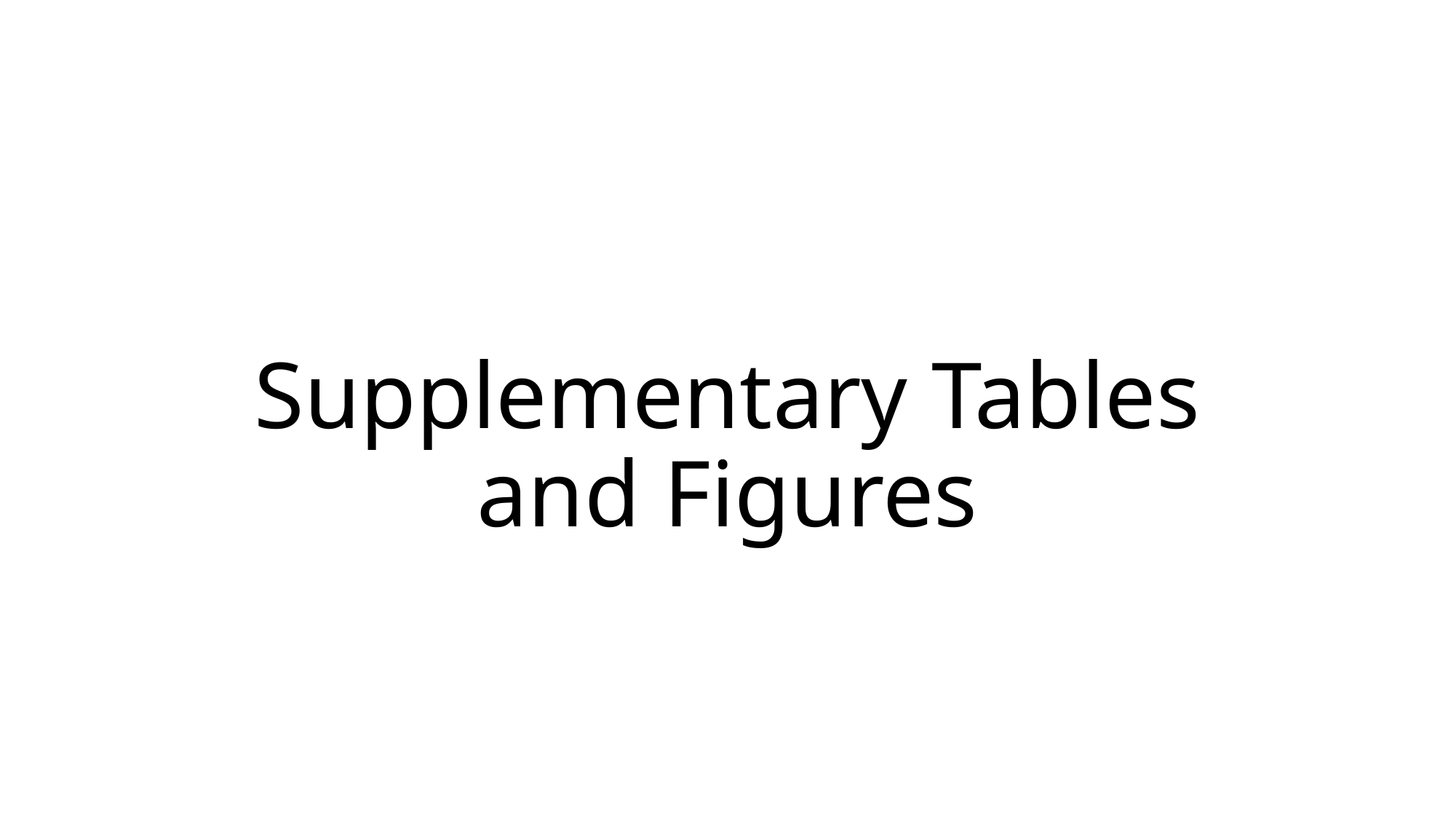

#### Supplementary Tables and Figures

### Slide 2
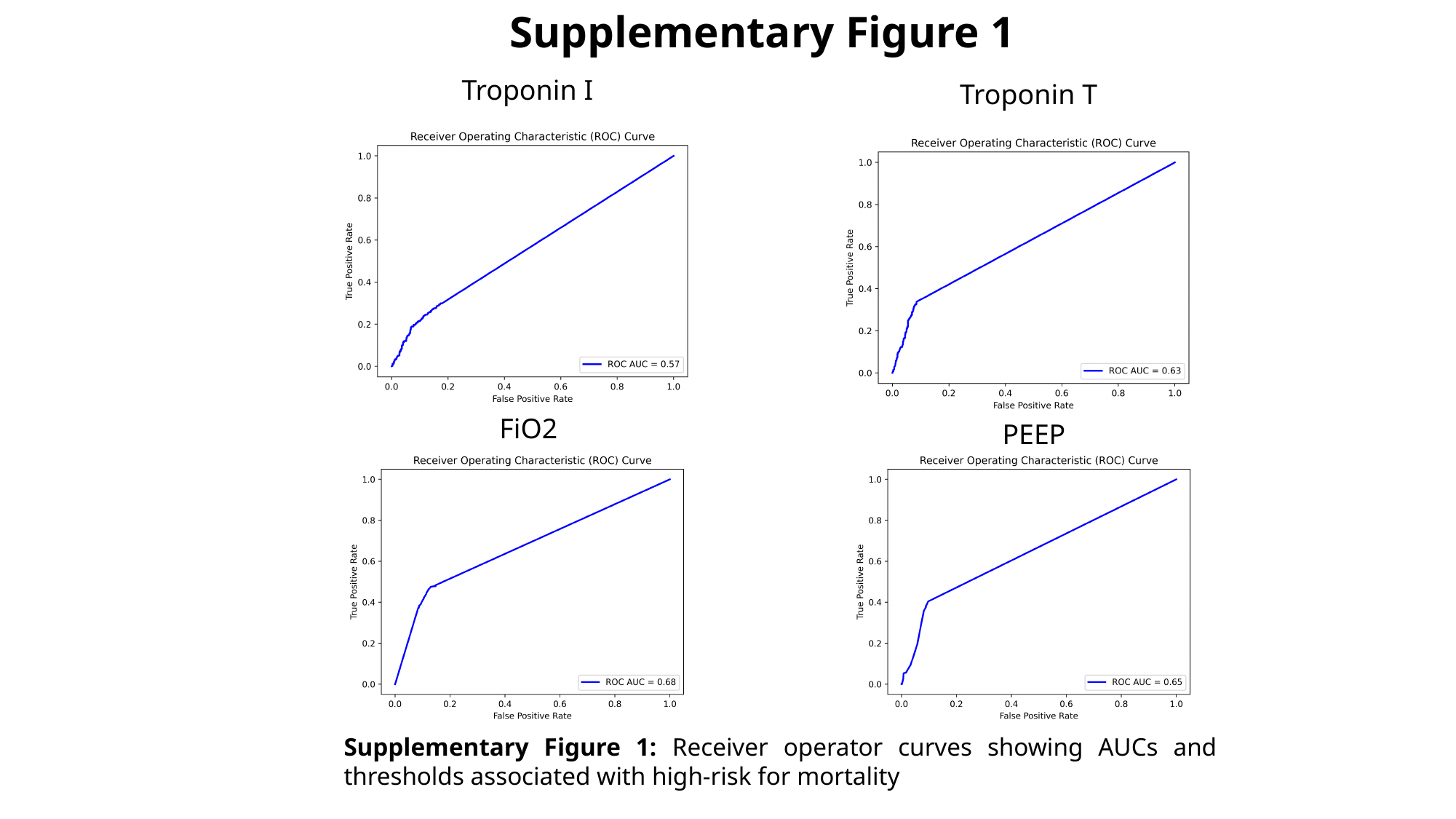

Supplementary Figure 1
Troponin I
Troponin T
FiO2
PEEP
Supplementary Figure 1: Receiver operator curves showing AUCs and thresholds associated with high-risk for mortality

### Slide 3
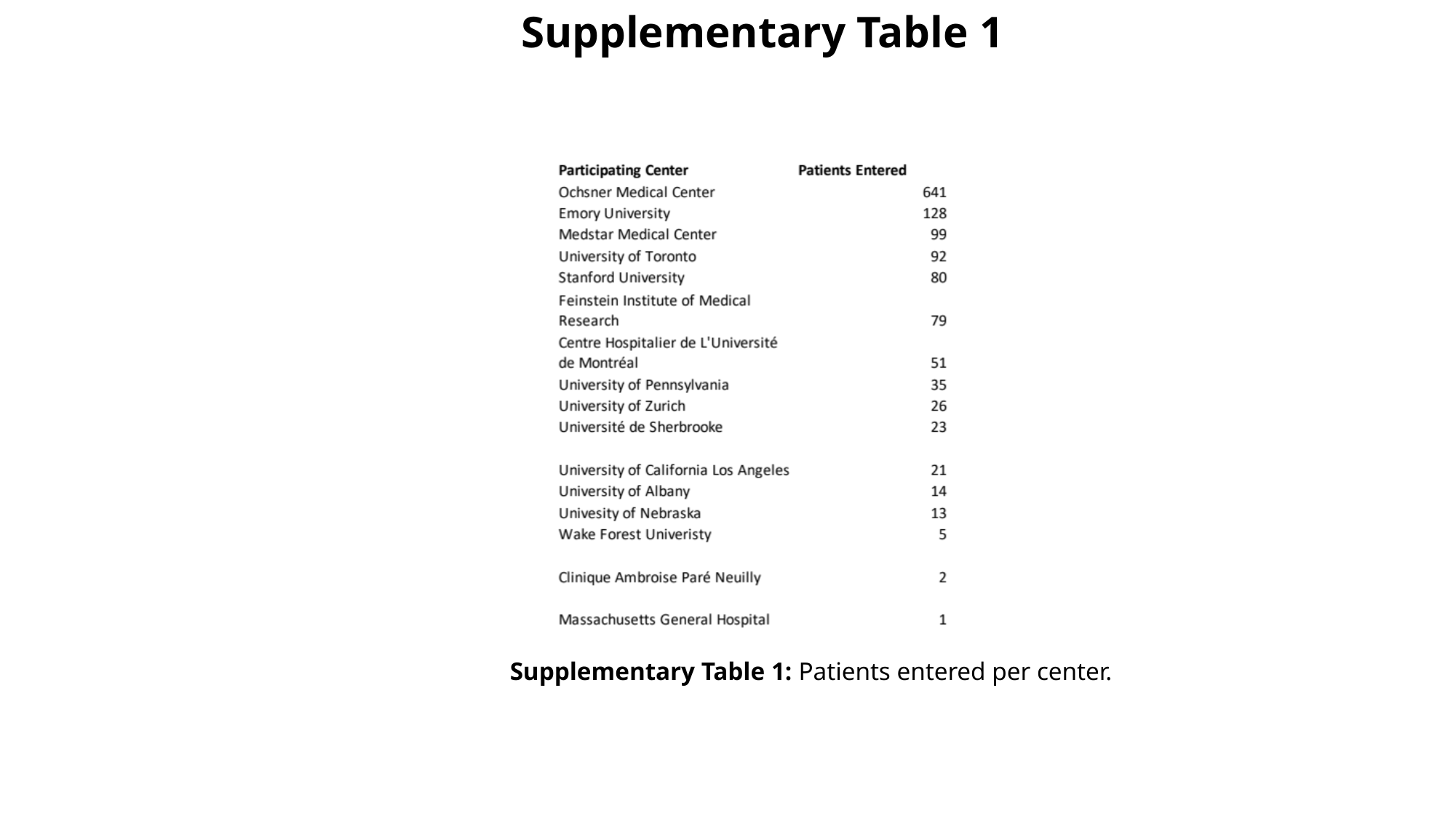

Supplementary Table 1
Supplementary Table 1: Patients entered per center.

### Slide 4
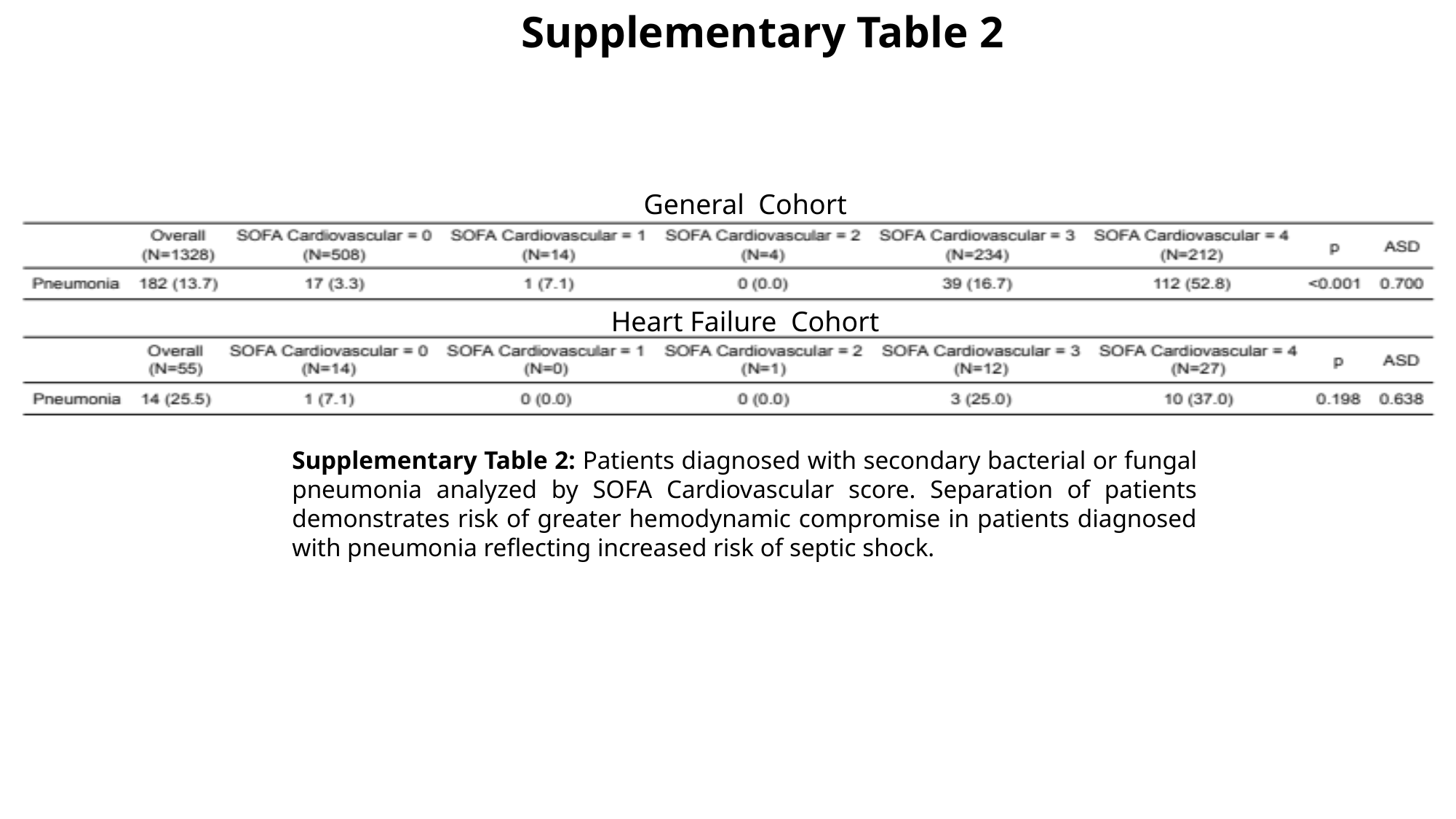

Supplementary Table 2
General Cohort
Heart Failure Cohort
Supplementary Table 2: Patients diagnosed with secondary bacterial or fungal pneumonia analyzed by SOFA Cardiovascular score. Separation of patients demonstrates risk of greater hemodynamic compromise in patients diagnosed with pneumonia reflecting increased risk of septic shock.
